# Wavelet analysis reveals non-stationary cardiovascular rhythms associated with delirium and deep sedation in ICU patients

**DOI:** 10.64898/2026.04.22.26351455

**Authors:** Jayanth Sreekanth, Eduardo Salgado-Baez, Andreas Edel, Elias Gruenewald, Sophie K. Piper, Claudia Spies, Felix Balzer, Sebastian Daniel Boie

## Abstract

Routine ICU data offers valuable insights into daily physiological rhythms. While traditional methods assume these cycles maintain fixed periods and amplitudes, their inherent variability requires dynamic estimation of instantaneous trends. Wavelet transform effectively resolves circadian oscillations, especially for frequently measured vital parameters. We present novel extensions to the Continuous Wavelet Transform (CWT) power spectral analysis to better detect and segment subtle temporal patterns. Using this approach, we uncover hidden circadian patterns in cardiovascular vitals such as Heart Rate (HR) and Mean Blood Pressure (MBP) measured over five days in a retrospective cohort of 855 ICU patients. By quantifying non-stationary rhythms, we identified diurnal and semi-diurnal oscillations varying in period and power according to delirium and deep sedation. Notably, HR exhibits a clear diurnal and semi-diurnal rhythm when delirium is absent. Overall, our framework supports the CWT as a powerful tool for analyzing complex physiological signals, particularly vital signs. Crucially, our findings suggest that cardiovascular rhythm disruption can be associated with ICU-related delirium and deep sedation.

## Introduction

The complex environment of an Intensive Care Unit (ICU), characterized by sedation, procedures, artificial noise and lighting, inadvertently disrupts patients’ sleep-wake cycle and circadian rhythm^1,2^. Circadian misalignment is associated with cardiovascular disease, cancer, response to infection, inflammation, disruption of glucose homeostasis, and psychiatric and neurodegenerative diseases, especially delirium^3–5^. Cardiovascular physiology is influenced by various mechanisms involving the autonomic nervous system and peripheral clocks^6^. Under normal conditions, these mechanisms produce a clear diurnal variation, aligning bodily functions with the sleep-wake cycle^6,7^. However, this adaptive ability decreases during critical illness, leading to a change in regular patterns^8–11^. Emerging evidence suggests that such circadian changes are closely connected to brain health, serving as early indicators of cognitive impairment and dementia^12–14^. Considering this complex relationship, examining periodic patterns in Blood Pressure (BP) and Heart Rate (HR) is an initial step to understanding the association between circadian disruptions and adverse cognitive conditions.

McKenna et al. highlight the finding of an optimal assessment tool as a key area of research in clinical chronobiology^15^. While some studies have applied peak-nadir excursion^10,11^ or cosinor regression^9,16,17^ to physiological signals, the dynamic nature of clinical routine data generated from continuous patient monitoring benefits from advanced signal processing methods to capture transient patterns. The wavelet transform has been valuable in studying the instantaneous trend of the circadian rhythm over time and holds significant potential for applications in the field of chronobiology and circadian medicine^18–25^. The wavelet transforms employ basis functions that are localized in both time and frequency with an effective window size that varies with scale. This multi-resolution property enables fine temporal resolution at high frequencies and fine frequency resolution at low frequencies, making wavelets particularly well suited for the analysis of non-stationary and multi-scale signals^26–30^. Therefore, to characterize dynamic periodicities and rhythm strength, the Continuous Wavelet Transform (CWT) is highly relevant for continuously monitored vital parameters at bedside.

The primary objective of this study is to develop and demonstrate a robust methodological framework using CWT to effectively uncover subtle periodic oscillations within vital parameters. By introducing an approach for tracking and segmenting maxima in wavelet power spectral analysis, we characterize transient dynamic properties of cardiovascular vitals associated with medical conditions. Our approach goes beyond conventional Fourier- and Cosinor-based methods that assume stationarity. Furthermore, we assess the clinical utility of our approach by applying it to multi-day HR and Mean Blood Pressure (MBP) recordings from 855 ICU patients, characterizing differences in cardiovascular patterns based on cognitive conditions such as with-delirium, without-delirium, and deep sedation.

## Results

An overview of the methods, including signal representation, is outlined in Figure 1. Here in the results section, we apply CWT and a new approach to identify and segment periodic components in clinical routine data, using HR of an individual as an example. Later, we present the results from applying this approach to a cohort, uncovering temporal patterns in HR and MBP associated with deep sedation and delirium.

**Figure 1.**
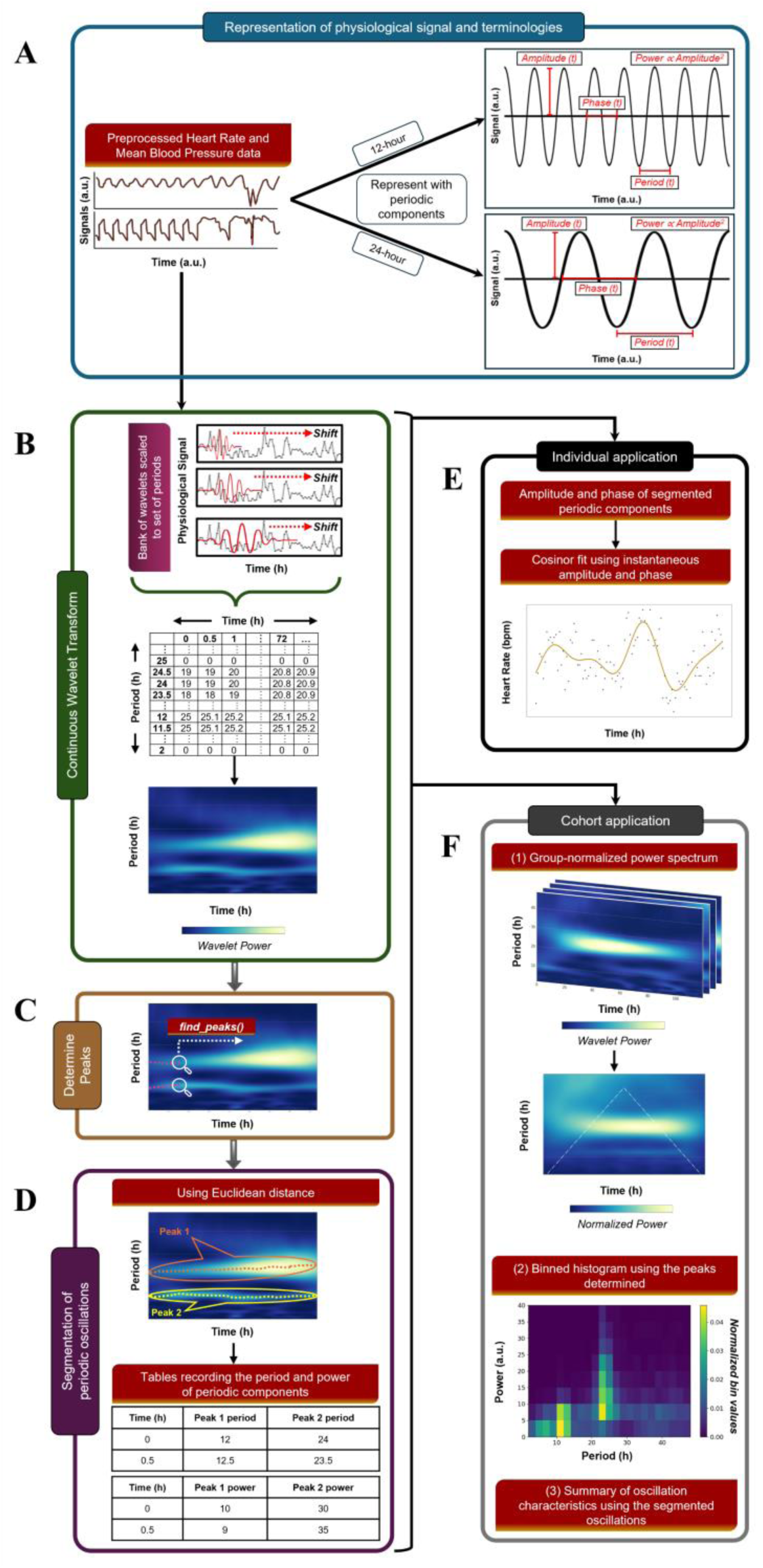
Graphical overview of our CWT-based method for analyzing periodic oscillations in physiological signals. The workflow is divided into conceptual representations, core methodological steps and practical applications: (A) Illustration of the breakdown of a physiological signal into multiple periodic components. It highlights key signal parameters - amplitude, phase and period - and includes that power is proportional to the square of the amplitude^31^. (B) Step 1 showing the application of CWT to the pre-processed physiological signal, yielding a comprehensive power spectrum. (C) Step 2 highlighting the identification of distinct peaks within the generated power spectrum. (D) Step 3 demonstrating the segmentation of periodic oscillations using their Euclidean distance and their corresponding period and power values recorded into data tables. (E) Provides a practical example of the steps applied to a single patient’s HR data, demonstrating how instantaneous amplitude and period extracted from the steps are utilized for cosinor fitting. (F) Application of the steps to a cohort of patients where distinct oscillation patterns emerge, allowing us to explore group-wise differences of periodic oscillations. Abbreviations: CWT, Continuous Wavelet Transform; HR, Heart Rate

### Continuous wavelet transform and cosinor regression applied to clinical routine data

Applying CWT on clinical routine time series yields a time-resolved power spectrum. Typically, in the power spectrum, multiple non-stationary components are found that can be tracked across time. These dynamic temporal changes are observed especially for multi-day recordings.

When using CWT to decompose a complex waveform into periodic components, one observes edge effects, which are due to the period-dependent finite size of the wavelet. Commonly, one defines a Cone of Influence (CoI) by demanding the wavelet amplitude envelope to decay to 1/𝑒. For the Morlet wavelet, the time that the envelope has decayed to 1/𝑒 (see methods) is given by

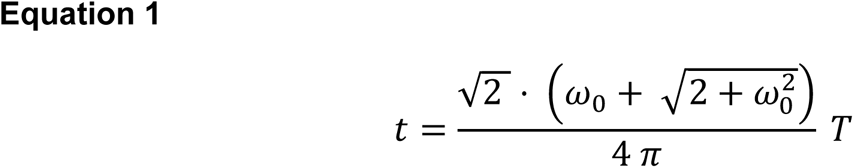

where 𝜔_0_is a constant (typically 𝜔_0_ = 2𝜋), and 𝑇 is the period of interest. Specifically, in the context of circadian medicine, where one may be interested in the period band between 22 – 26 hours, period and power estimates in the first 37.2 h (corresponding to T = 26 h) are affected by the edge effects (cf. Figure 2). The edge effects are symmetrical and occur at both the beginning and the end of the time series. Importantly, if periodic components are examined within a fixed window (e.g., the first 5 days), the CoI extends beyond this window when additional data exist but is not excluded before analysis.

**Figure 2.**
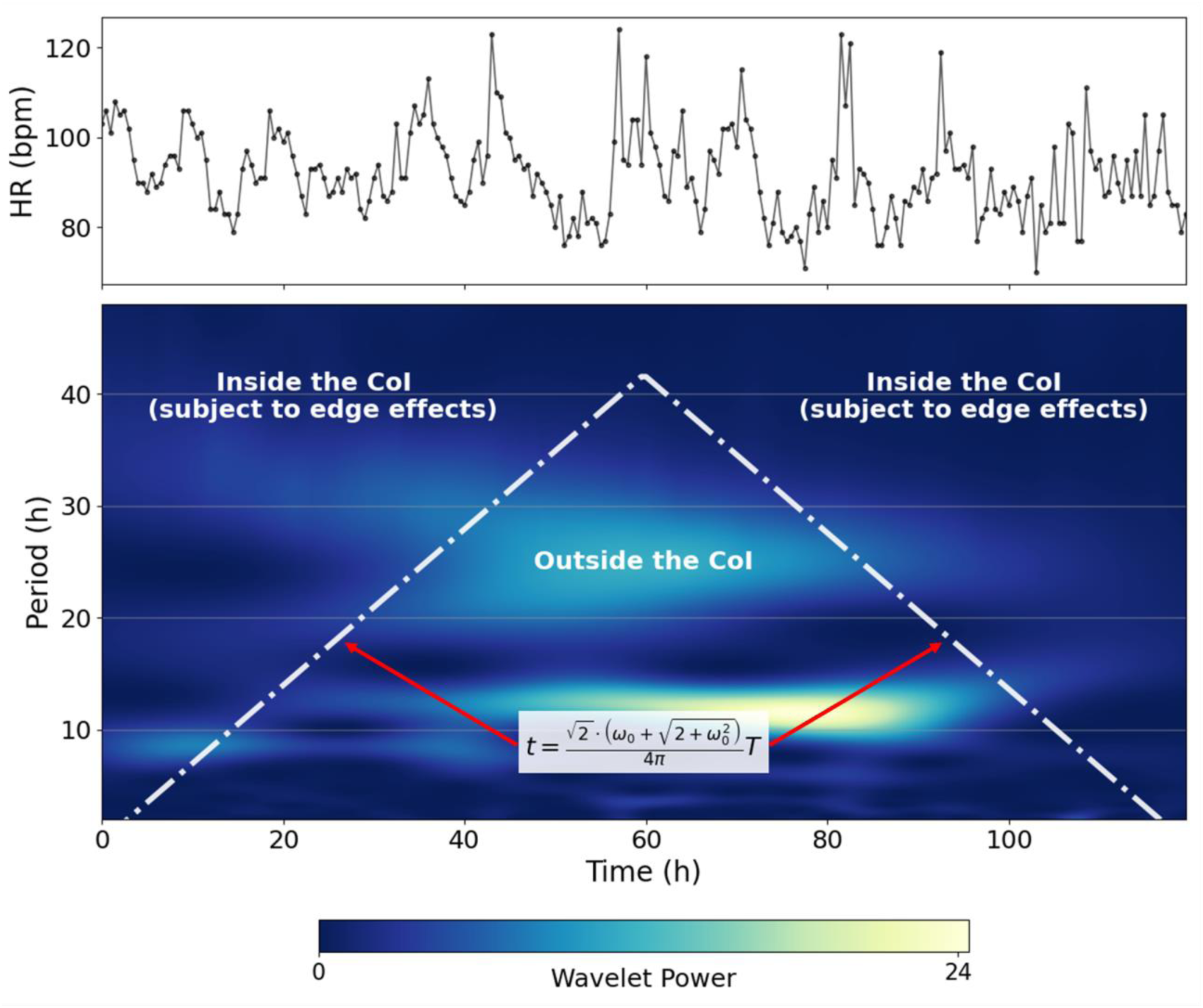
Wavelet power spectrum for a 120-hour (5-day) measurement period, showing the CoI boundary to account for edge effects. Measurements above the boundary are inside the CoI and subject to edge effects; measurements below the boundary are outside the CoI and not subject to edge effects. The parameters 𝝎_𝟎_ is a constant, usually assigned a value of 𝟐𝝅, and 𝑻 represents the defined period of interest. Abbreviations: h, hours; CoI, Cone of Influence

After fitting the cosinor curves to the HR data, the one-component cosinor model yields an R^2^ (coefficient of determination) = 0.21, while the two-component model improved the fit to R^2^ = 0.52. A cosinor model incorporating non-stationary amplitude and phase derived from CWT yields an R^2^ = 0.58 (Figure 3B). In this model, the number of components is not constant, as periodic components can appear or disappear as observed in the spectrogram. Consistent with this description, the model fit exhibits sharp transitions, attributable to the presence or absence of periodic components.

**Figure 3.**
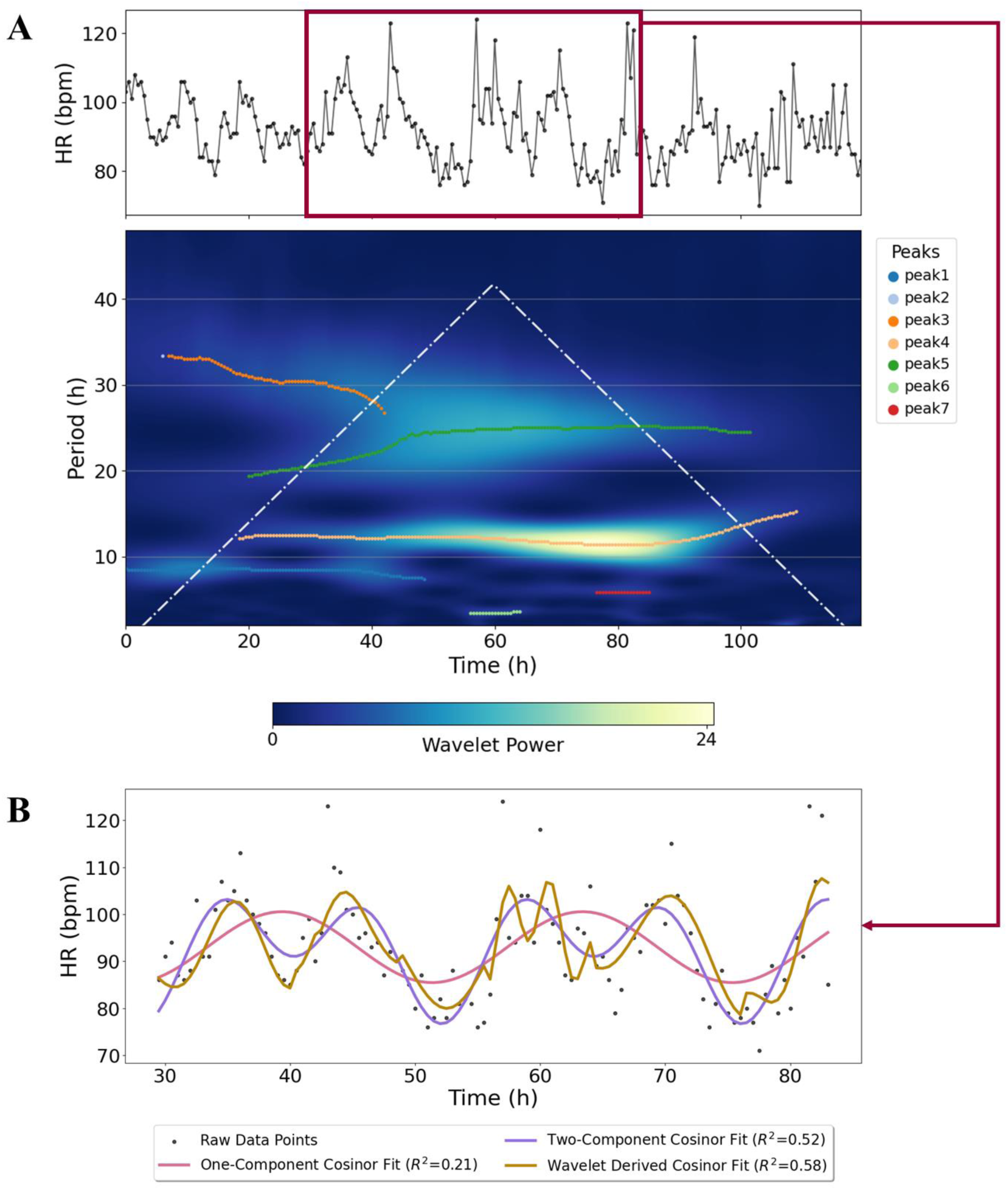
Identification and segmentation of non-stationary periodic components in an individual patient signal using CWT, followed by the cosinor model fits. *(A)* Multi-panel visualization showing a complex, non-stationary HR signal at the top, representing the raw data of an individual patient, and a power spectrum displaying multiple color-coded periodic components, at the bottom. Each color-coded peak represents a distinct periodic component, such as semidiurnal (around 12h) and diurnal (around 24h). Dashed white lines in the spectrum refer to the CoI. *(B)* Multiple cosinor models fitted to the raw HR data of an individual patient using timestamps not influenced by the edge effects (ca. 29.5 – 83 hours). The displayed cosinor fits correspond to: a one-component model with a constant period of 24-h (red-violet curve), a two-component model with a constant period of 12-h and 24-h (medium-purple curve), and a multi-component model with wavelet-derived instantaneous amplitude and phase (dark-gold curve). Abbreviations: HR, Heart Rate; bpm, beats per minute; h, hours; CWT, Continuous Wavelet Transform; CoI, Cone of Influence

### Cohort description and baseline characteristics

We apply CWT on a cohort to reveal dynamic properties of HR and MBP for different groups. The study cohort comprises 2,433 cases who tested positive for SARS-CoV-2 and were hospitalized between January 2020 and February 2022. Of these, 2,362 unique patients required ICU treatment. Among them, 1,469 patients had measurements spanning at least five consecutive days, of which 1,419 had measurements recorded within the first three weeks after admission. Ultimately, the final analysis included 855 patients with consistent delirium or sedation status classification across all five measurement days, based on CAM-ICU, patient chart review, or RASS. The flow diagram of patient selection is described in Figure 4.

**Figure 4.**
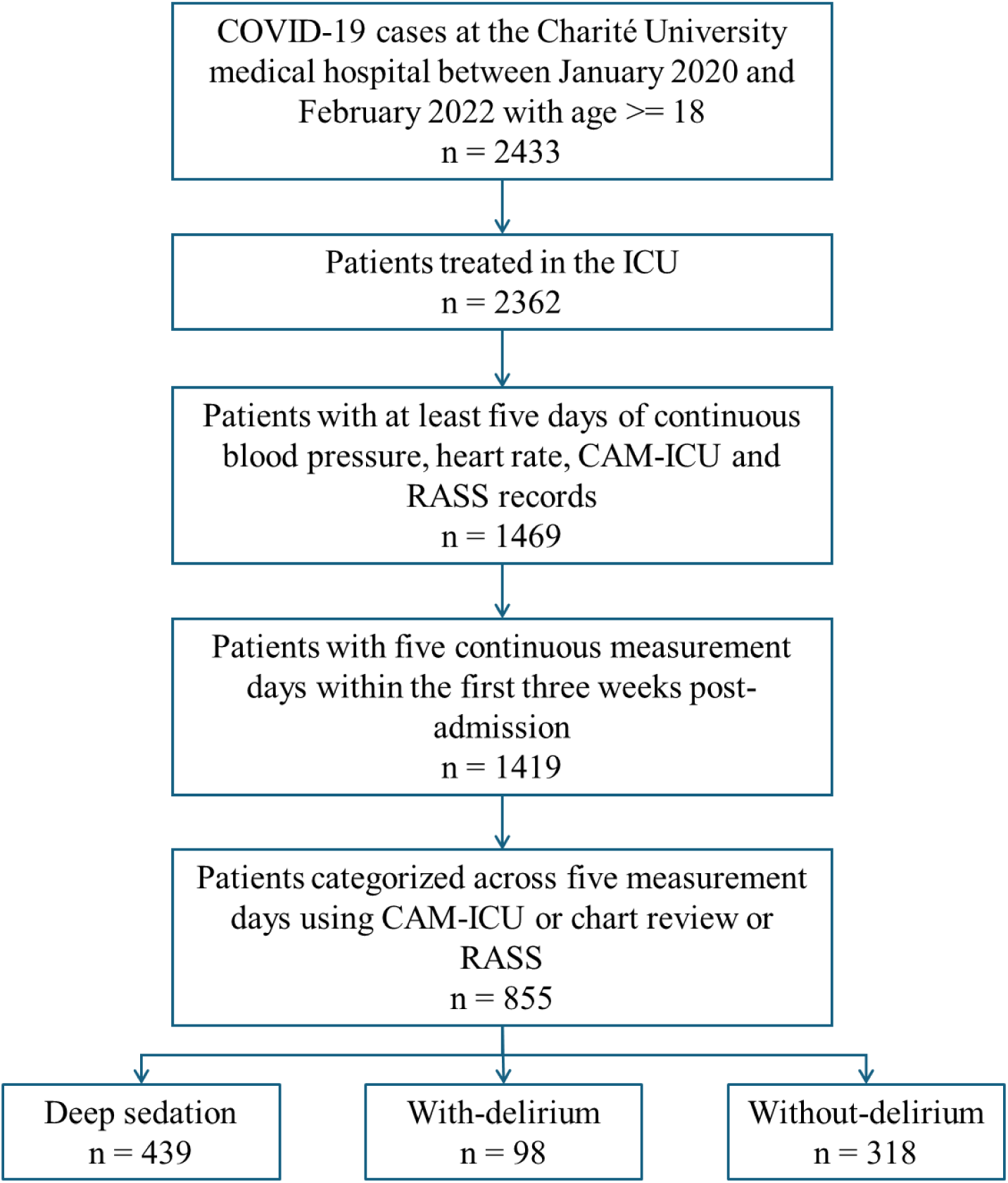
Patient selection and stratification process. Outline of patient inclusion with counts for each step, from initial screening to the classification of the groups. Abbreviations: COVID-19, Coronavirus disease 2019; ICU, Intensive Care Unit; CAM-ICU, Confusion Assessment Method for the Intensive Care Unit; RASS, Richmond Agitation-Sedation Scale

The basic characteristics of the 855 patients are summarized in Table 1, stratified by delirium or sedation status across five consecutive measurement days. The median hospital stay was approximately 17 days longer in the deep sedation group compared with the other two groups, and a substantially greater proportion of this time (88.9%) was spent in the ICU. The overall cohort exhibited a male-to-female ratio of 2.3:1 (69.7% vs 30.3%), and each group similarly consisted of more than 66% male patients. Hospital mortality rates differed markedly between the groups: only 18.9% of patients without delirium died, whereas 51% of deep sedated patients died.

**Table 1.** Summary of the demographic and clinical characteristics of the patient cohort. Categorical variables are presented as count (percentage) and continuous variables as median [Q1, Q3]. Abbreviations: BMI, Body Mass Index; ICU, Intensive Care Unit; Q1, first quartile; Q3, third quartile; CAM-ICU, Confusion Assessment Method for the Intensive Care Unit; GCS, Glasgow Coma Scale

|  | Missing | Overall | Deep sedation | With-Delirium | Without-Delirium |
| --- | --- | --- | --- | --- | --- |
| **n** |  | 855 | 439 | 98 | 318 |
| Hospital duration in days | 0 | 25 [14,45] | 36 [20,60] | 20 [14,33] | 18 [12,31] |
| ICU duration in days | 0 | 17 [10,38] | 32 [19,52] | 12 [8,17] | 10 [7,15] |
| Age | 0 | 61.0<br>[52.0,69.0] | 59.0<br>[50.0,65.0] | 73.5<br>[64.2,81.8] | 62.0<br>[53.0,71.0] |
| Sex (male) |  | 596 (69.7) | 319 (72.7) | 66 (67.3) | 211 (66.4) |
| BMI | 181 | 27.9<br>[24.7,32.7] | 29.4<br>[26.0,34.3] | 24.9<br>[22.9,27.8] | 27.1<br>[24.4,31.3] |
| Hospital mortality |  | 317 (37.1) | 224 (51.0) | 33 (33.7) | 60 (18.9) |
| Hospital days with "positive" CAM-ICU |  | 0 [0,2] | 0 [0,3] | 2 [1,5] | 0 [0,0] |
| Hospital days with "unknown" CAM-ICU |  | 10 [0,26] | 23 [14,36] | 2 [1,5] | 0 [0,1] |
| Hospital days with “negative” CAM-ICU |  | 7 [1,12] | 1 [0,11] | 6 [3,10] | 9 [7,13] |
| GCS on hospital discharge | 432 | 4 [3,15] | 3 [3,14] | 7 [3,13] | 11 [3,15] |
| **Circulation system disorders** |  |  |  |  |  |
| Atherosclerosis |  | 25 (2.9) | 6 (1.4) | 6 (6.1) | 13 (4.1) |
| Chronic ischaemic heart disease |  | 129 (15.1) | 46 (10.5) | 30 (30.6) | 53 (16.7) |
| Heart failure |  | 192 (22.5) | 104 (23.7) | 30 (30.6) | 58 (18.2) |
| Essential (primary) hypertension |  | 479 (56.0) | 235 (53.5) | 64 (65.3) | 180 (56.6) |
| Hypertensive heart disease |  | 50 (5.8) | 25 (5.7) | 7 (7.1) | 18 (5.7) |
| Hypertensive renal disease |  | 4 (0.5) | 1 (0.2) | 0 (0.0) | 3 (0.9) |
| Hypertensive heart and renal disease |  | 2 (0.2) | 0 (0.0) | 0 (0.0) | 2 (0.6) |
| **Mental and behavioral disorders** |  |  |  |  |  |
| Depressive episode |  | 28 (3.3) | 12 (2.7) | 6 (6.1) | 10 (3.1) |
| Recurrent depressive disorder |  | 8 (0.9) | 4 (0.9) | 1 (1.0) | 3 (0.9) |
| Bipolar affective disorder |  | 2 (0.2) | 2 (0.5) | 0 (0.0) | 0 (0.0) |
| Schizophrenia |  | 8 (0.9) | 5 (1.1) | 2 (2.0) | 1 (0.3) |
| Other anxiety disorders |  | 57 (6.7) | 34 (7.7) | 7 (7.1) | 16 (5.0) |
| **Neurological disorders** |  |  |  |  |  |
| Alzheimer disease |  | 4 (0.5) | 1 (0.2) | 3 (3.1) | 0 (0.0) |
| Parkinson disease |  | 7 (0.8) | 2 (0.5) | 1 (1.0) | 4 (1.3) |
| Multiple sclerosis |  | 2 (0.2) | 2 (0.5) | 0 (0.0) | 0 (0.0) |

The median Glasgow Coma Scale (GCS) score upon hospital discharge indicated varying levels of consciousness: 3 (most severely impaired) for the deep sedation, 7 (severely impaired) for the with-delirium and 11 (moderately impaired) for the without-delirium groups. The presence of circulation system disorders was found to be high in the group of patients with delirium. Hypertension was nearly equal among the groups, with an overall prevalence of 56%, except for the delirious group, being 65%. The patient group with delirium exhibited higher chronic ischaemic heart disease (n=30; 30.6%) compared to the deep sedation group (n=46; 10.5%) and the group without delirium (n=53; 16.7%). Neurological disorders were diagnosed in fewer than 1% of patients; therefore, their direct influence as confounders on the cognitive outcomes was considered minimal. A small proportion of the cohort presented mental and behavioral disorders (<7%). An extended table of clinical parameters is provided in Supplementary Table 1.

The summary of clinical characteristics measured over five measurement days is in Supplementary Table 2. Only 218 patients (25.5%) out of 855 required imputations for HR and MBP measurements, and the overall median for the timestamps imputed was 10 (5 hours due to measurements every 30 minutes). In the deep sedation group, morphine was administered for a median of 13 minutes (range: 0–5,761). In contrast, the median duration of intravenous systemic sufentanil administration varied significantly by cognitive condition: 55 minutes (range: 0–5,161) for patients with delirium versus 231 minutes (range: 0–5,867) for those in deep sedation. Independently, patients with delirium received regional epidural anesthesia (ropivacaine and sufentanil) for a median of 11 minutes (range: 0–1,047). Besides these, deep sedated patients received more vasopressors and sedatives than the other two groups. Prognostic scores reflected the severity of illness and delirium or sedation status across the five measurement days. The deep sedated group demonstrated a median SOFA score of 11 [10–12], indicating severe organ dysfunction, accompanied by a high APACHE II score of 26 [16–32] and the greatest need for therapeutic intervention (TISS-10: 14 [10–17]).

### Wavelet Analysis reveals different group-wise patterns in physiological signals

Vital parameter signals, such as HR and MBP, were continuously recorded in the ICU every 30 minutes in 855 patients. After applying CWT, the group-stratified power spectrum in Figure 5 reveals periodic oscillations in the dynamics of the vital signs over a five-day observation window.

**Figure 5.**
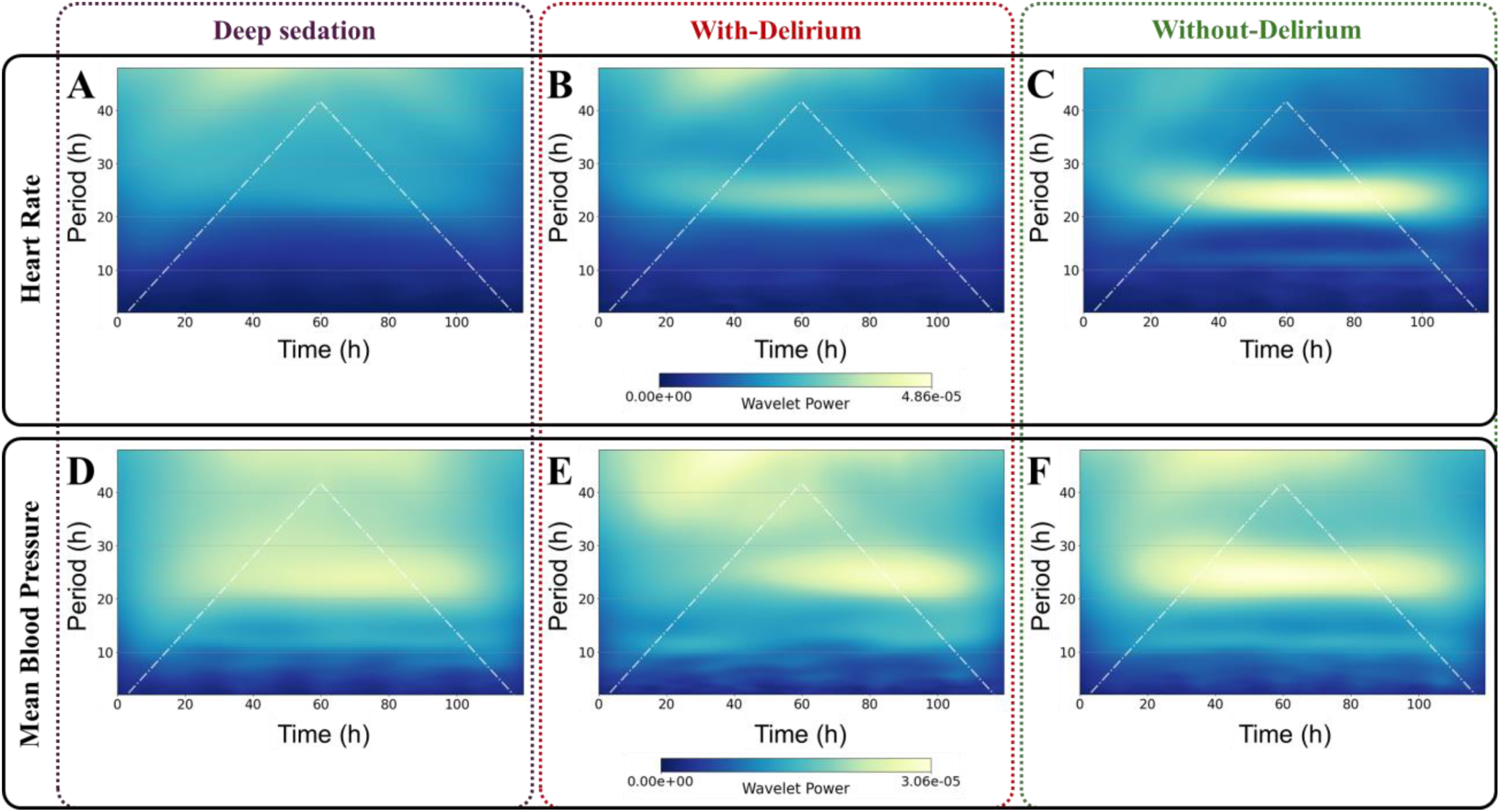
Group-normalized CWT power spectrum of HR and MBP. The color bar of the spectrum indicates group-normalized power of the signal variations. The white lines refer to the CoI. Plots *(A-C)* show the power spectrum for HR, and plots *(D-F)* for MBP. The columns show deep sedation *(A, D)*, with-delirium *(B, E),* and without-delirium groups *(C, F)*. Abbreviations: HR, Heart Rate; MBP, Mean Blood Pressure; h, hours; CWT, Continuous Wavelet Transform; CoI, Cone of Influence

The CWT spectrograms of the HR signal (Figure 5A, B, C) show power differences across different periods for different groups. Periodic components of the HR signal are unclear and substantially attenuated in deep sedated patients (Figure 5A). Patients with delirium exhibit minor oscillations with a period of approximately 22–26 hours (Figure 5B). A strong and persistent oscillation is observed between t = 35 and 90 hours at a period of 22–26 hours in the without-delirium group (Figure 5C). This dominant cycle is accompanied by a weaker but persistent semidiurnal oscillation (period ∼12 hours) that extends across a broader time window (20–100 hours).

MBP variations display a broad spectrum of oscillatory periods (Figure 5D, 5E, 5F). This pattern is particularly evident in deep sedated patients, in whom MBP oscillations—including both diurnal and semidiurnal cycles—are apparent even when HR oscillations are not (Figure 5D and 5A). In patients with delirium, MBP oscillations with a period of approximately 22–26 hours emerge from t = 48 hours onward, whereas the semidiurnal component remains unclear (Figure 5E). In patients without delirium, the MBP fluctuation pattern closely mirrors the HR pattern (Figure 5F and 5C), showing both diurnal and semidiurnal oscillations; however, the diurnal MBP period appears broader.

The spectrograms in Figure 5 demonstrate clear group-wise differences in the strength and temporal consistency of cardiovascular rhythms, with markedly reduced power and stability of the semidiurnal and diurnal oscillations in both the with-delirium and deep sedation groups.

### Characteristics of group-wise dominant and non-dominant oscillations

The peaks identified in the spectrograms correspond to periodic oscillations. By utilizing the period, power and timing of the peaks derived from the spectrum, a comparison of bivariate histograms—constructed with predefined bin widths—highlights a clear shift in the distribution of peaks in the spectrograms of HR across three groups (Figure 6). The power–period histograms of the peaks reveal the presence of multiple periodic components, with increased power observed around 12-hour (semidiurnal) and 24-hour (diurnal) periods. These periodic oscillations are especially pronounced in the group without delirium compared to the group with delirium (Figure 6B, 6C), whereas no distinct spectral power can be observed across any periods in the deep sedation group (Figure 6A). The period–time histograms show that these periodic oscillations remain stable and persistent over time in patients without delirium but are less robust and more fragmented in those with delirium (Figure 6E, 6F).

**Figure 6.**
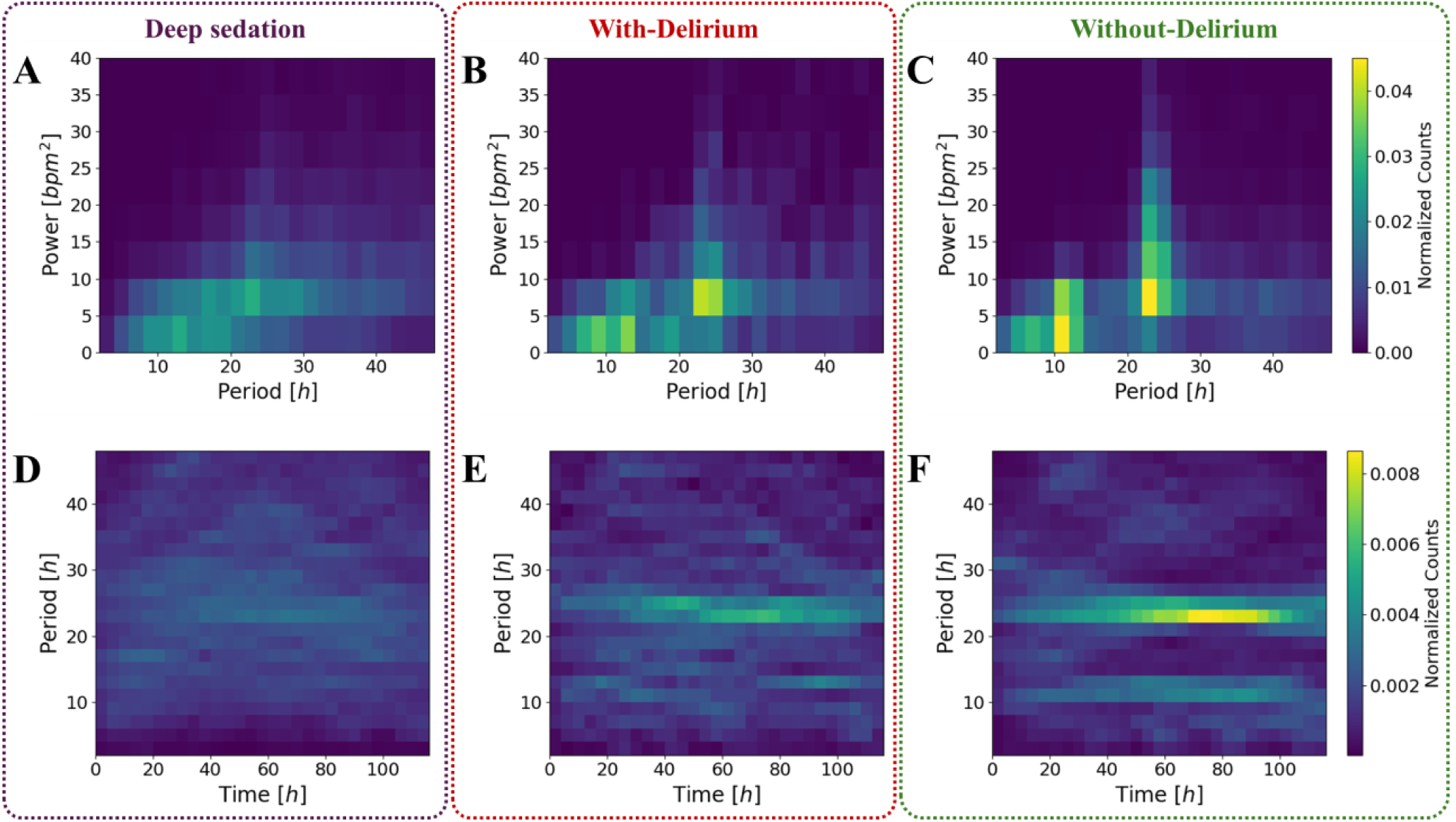
Bi-dimensional histograms with arbitrary bin widths showing the normalized joint distribution of oscillations in HR measurements in deep sedation, with-delirium and without-delirium groups. The histograms include a joint distribution between the period in hours (x-axis) and power in *bpm*^2^ (y-axis) of oscillations in the *(A)* deep sedation, *(B)* with-delirium and *(C)* without-delirium group. Hourly temporal changes of oscillation period in hours in the *(D)* deep sedation, *(E)* with-delirium and *(F)* without-delirium groups are shown. Abbreviations: HR, Heart Rate; bpm, beats per minute; h, hours

In contrast to the clear group distinction observed in HR dynamics over multiple days, MBP exhibits a broad and overlapping distribution of periodic oscillations across all groups, suggesting a high degree of inconsistent variability, especially in the deep sedation and with-delirium groups. Whereas diurnal and semidiurnal periodicities remain observable in the without-delirium group (Supplementary Figure 2). The bivariate distribution of individual peaks (Figure 6 and Supplementary Figure 2) shows strong correspondence with the features observed in the group spectrum (Figure 5).

### Segmentation and group-wise summarization of periodic components in the Spectrum

The periodic oscillations identified in the power spectrum, once segmented, become distinct periodic components in the physiological signal. The analysis of separated periodic components in the HR spectrum in the region unaffected by the CoI reveals parameters of between-group differences (Table 2). For the 10 – 14 hours periodic component, significant differences (p-adjusted < 0.05) were observed between the deep sedation and without-delirium groups across all reported parameters, except for the oscillation start time and the average power. In contrast, no statistically significant differences were found across any parameter when comparing deep sedation vs. with-delirium groups or with-delirium vs without-delirium groups. For the 22 – 26 hours periodic component, no significant differences were seen between the deep sedation and with-delirium groups, except for the number of patients who didn’t have oscillations within this specific period or had oscillations inside the CoI. However, significant differences were noted between the deep sedation and without-delirium groups for all parameters, and between the with-delirium and without-delirium groups for all parameters except the oscillation start time and the number of patients who didn’t have oscillations within this specific period or had oscillations inside the CoI. These differences in the number and magnitude of affected parameters align with the group differences visually apparent in the bivariate histograms (Figure 6).

**Table 2.** Pair-wise statistical differences of group summarized periodic components in HR oscillations. All continuous variables are presented as median [Q1, Q3] and categorical variables as count (percentage). Group differences were evaluated using the Kruskal-Wallis test (continuous) and the Chi-squared test (categorical). P-values were adjusted using the Bonferroni correction for each set of comparisons with the correction factor of n=14. The patients excluded are the ones who didn’t have oscillations within the specific period or had specific periodic oscillations inside the CoI. Abbreviations: HR, Heart Rate; h, hours; bpm, beats per minute; N.S., Not Significant; Q1, first quartile; Q3, third quartile; CoI, Cone of Influence

| Oscillation Characteristics | Oscillation Period (h) | Overall | Deep sedation | With-Delirium | Without-Delirium | P-Value (adjusted) [Deep sedation vs With-Delirium] | P-Value (adjusted) [Deep sedation vs Without-Delirium] | P-Value (adjusted) [With-Delirium vs Without-Delirium] | P-Value (adjusted) [Overall] |
| --- | --- | --- | --- | --- | --- | --- | --- | --- | --- |
| **n** |  | 855 | 439 | 98 | 318 |  |  |  |  |
| Average duration of oscillations (*h*) | **10 - 14** | 17 [8,30] | 14 [6,24] | 16 [9,27] | 22 [11,36] | N.S. | <0.001 | 0.098 | <0.001 |
| Minimum oscillation duration (*h*) |  | 12 [4,27] | 10 [4,22] | 11 [4,22] | 16 [6,35] | N.S. | <0.001 | 0.413 | 0.003 |
| Maximum oscillation duration (*h*) |  | 20 [10,34] | 18 [7,29] | 19 [10,33] | 27 [13,42] | N.S. | <0.001 | 0.083 | <0.001 |
| Oscillation start hour |  | 36.5 [19.0,61.5] | 37.5 [19.5,63.2] | 31.8 [18.5,57.2] | 36.5 [18.5,59.0] | N.S. | N.S. | N.S. | N.S. |
| Oscillation end hour |  | 87.0 [59.5,101.0] | 79.0 [51.5,99.5] | 89.0 [47.4,99.6] | 93.5 [69.6,101.5] | N.S. | <0.001 | 0.169 | <0.001 |
| Average power (*bpm*<sup>2</sup>) |  | 5 [4,6] | 4 [4,6] | 4 [3,5] | 5 [4,6] | N.S. | N.S. | 0.158 | 0.579 |
| Count of patients excluded (no peaks or all peaks inside Col) |  | 306 (35.8) | 192 (43.7) | 38 (38.8) | 76 (23.9) | N.S. | <0.001 | 0.082 | <0.001 |
| Average duration of oscillations (*h*) | **22 - 26** | 29 [13,48] | 20 [8,36] | 24 [11,48] | 40 [22,50] | N.S. | <0.001 | 0.005 | <0.001 |
| Minimum oscillation duration (*h*) |  | 29 [11,48] | 20 [8,36] | 24 [10,48] | 40 [21,50] | N.S. | <0.001 | 0.013 | <0.001 |
| Maximum oscillation duration (*h*) |  | 30 [14,48] | 20 [8,36] | 25 [11,48] | 40 [24,50] | 0.901 | <0.001 | 0.004 | <0.001 |
| Oscillation start hour |  | 37.0 [35.0,54.5] | 39.0 [35.0,61.0] | 37.2 [35.0,54.5] | 36.5 [34.5,49.5] | N.S. | 0.046 | N.S. | 0.172 |
| Oscillation end hour |  | 83.8 [71.5,85.5] | 79.5 [62.5,85.0] | 83.5 [67.2,85.5] | 84.5 [83.0,86.0] | N.S. | <0.001 | 0.025 | <0.001 |
| Average power (*bpm*<sup>2</sup>) |  | 10 [6,17] | 8 [4,13] | 10 [5,14] | 12 [7,19] | N.S. | <0.001 | 0.015 | <0.001 |
| Count of patients excluded (no peaks or all peaks inside Col) |  | 291 (34.0) | 206 (46.9) | 26 (26.5) | 59 (18.6) | 0.005 | <0.001 | N.S. | <0.001 |

Despite visual differences in the bivariate histograms of MBP (Supplementary Figure 2), the statistical summary of segmented peaks (periodic oscillations) reveals no significant inter-group differences for any periodic components (Supplementary Table 4), unlike the findings for HR.

## Discussion

In this study, we show that CWT provides a powerful framework for discovering dynamic, non-stationary oscillations in clinical routine data and for characterizing physiological periodicity in critically ill patients. We show that CWT applied to individual multi-day recordings can capture time-varying periodic components that are easily missed by other methods relying on assumptions of stationarity, which are rarely met in multi-day routine data^16,32–35^.

Conventional approaches, such as single- or multi-component cosinor models, assume fixed periodicities and therefore perform poorly when rhythms drift over time ^32,34^. Similarly, Fourier-based methods decompose a signal into global harmonics and do not preserve the timing of transient changes^32,34^. The CWT, by contrast, provides a mathematically well-defined time–frequency representation that accommodates drifting periods and transient oscillations^21,36^. Although empirical mode decomposition is another option for analyzing non-stationary physiological signals^37^, it is noise-sensitive and lacks a clear frequency interpretation^38,39^. The CWT, therefore, offers a more interpretable and robust approach for long ICU recordings, with potential direct application at the bedside^23,24,40^. Analogous to the sequential workflow published by Cunningham et al., in which dominant harmonics are first verified using a Lomb-Scargle periodogram prior to the cosinor model^41^, CWT can also be used as a precursor to obtain more realistic fits to routine data. This integration of CWT-derived parameters—specifically instantaneous amplitudes and periods—provides a dynamic, data-driven parameterization for cosinor analysis.

With our novel segmentation approach, we identified distinct group-wise differences in cardiovascular rhythmicity within a large cohort of ICU patients with Coronavirus disease 2019 (COVID-19). Patients without delirium showed strong and persistent diurnal (22–26 h) and semidiurnal (10-14 h) HR oscillations, whereas these rhythms are markedly attenuated or absent in deep sedated patients and substantially weakened in those with delirium. Statistical analysis of segmented HR oscillations illustrates significant differences in both diurnal and semidiurnal characteristics in deep sedated and delirious patients in comparison to those without delirium. MBP recordings show broader and less discriminative oscillatory patterns across groups. As the power of the CWT is a direct measure of variability in the signal^31^, a decrease in HR power in the delirium group compared to the without-delirium group suggests reduced variability, while very fragmented and low power seen in the deep sedation group indicates severe loss of variability. Given that BP variability is known to be reduced in non-dipping hypertensive patients^42^ and considering a significant portion of our cohort was hypertensive, we postulate that the decreased MBP power observed in the delirium and deep sedation group (Supplementary Figure 2) reflects a non-dipping phenotype.

Across all groups, HR time series displayed drifting amplitudes and periods, abrupt transitions in oscillatory structure, and changes in spectral power over multi-day windows. These fluctuations likely reflect a combination of intrinsic circadian and ultradian regulation, external environmental factors (e.g., sedation, light exposure, mechanical ventilation)^43^, and critical illness-associated organ and autonomic dysfunction^7,44^. Our findings align with the literature showing associations between circadian disruption in physiological signals and adverse outcomes, e.g., increased mortality with disruption of BP^7,12^, core body temperature in trauma patients^45^, temperature in critical illness myopathy^46^, and vitals such as respiratory rate, SpO2, and HR^47^. Additionally, core body temperature is thought to be a key driver in the circadian regulation of HR^48^.

The synchronization of cardiovascular physiology is intrinsically linked to brain health, where dysregulation of diurnal patterns serves as both a marker and a mechanism for cognitive impairment^13,14,49^. Thus, the loss of diurnal and semidiurnal structure in HR dynamics here may reflect underlying dysregulation of autonomic or neurophysiological systems in deep sedated patients. Specifically, deep sedation with agents like propofol causes parasympathetic dominance^50^, and acts as a myocardial depressant^51^, which, when coupled with simultaneous use of Esketamine and Ketamine, and prolonged vasopressor support with Norepinephrine, results in a complex but intensely managed suppression of intrinsic variability in the autonomic system^52^.

The dysfunction of the autonomic system extends to BP regulation because critically ill and sedated patients are unable to maintain a synchronized periodic cycle, resulting in a non-dipping pattern^53^. The observed broad, fragmented or shifted diurnal and semi-diurnal MBP oscillations confirm this severe disturbance. This loss of normal BP rhythm is critical for the brain, as non-dipping and reverse-dipping are associated with delirium^12^, cerebral small vessel disease^54^, subclinical brain damage^55^, and a nearly two-fold increased risk of Alzheimer’s disease^7,56^. These findings reinforce the importance of circadian and ultradian monitoring for early identification or risk stratification of delirium in the ICU and further confirm the association between circadian disruption and adverse outcomes.

Our framework opens the possibility for studying the transient effects of medications such as sedatives, vasopressors, or corticosteroids on cardiovascular rhythmicity over multiple days. Since hypertension alters baseline rhythmicity^42^, it is necessary to not only compare periodic patterns between survivors and non-survivors but specifically investigate whether patients across these three groups (deep sedation, with-delirium and without-delirium) exhibit distinct non-dipping and reverse-dipping profiles, as these patterns could provide additional yet critical insights into their pathophysiology^7^. Additionally, we plan to explore whether specific pharmacological, nutritional or environmental interventions (including controlled light exposure) can restore circadian or ultradian dynamics in critically ill patients.

This study has several limitations. First, although wavelet-based methods offer powerful tools for analyzing nonstationary physiological signals, their performance depends heavily on the choice of wavelet function^33^. The Morlet wavelet transform, widely used in biomedical signal analysis, may not be optimal in all situations, and inappropriate selection can lead to suboptimal feature extraction^20^. Second, challenges arise from irregular sampling, missing data, and artifacts common in ICU physiological recordings, as these factors may distort the spectral estimates and limit the precision of temporal–frequency interpretations.

Third, the observational design of the study precludes any statements about causality. Relationships identified between physiological patterns and clinical events cannot be interpreted as causal mechanisms. While cardiovascular physiology relies on timed and synchronized processes^57,58^, aging presents a potential confounder-particularly given that the median age of the with-delirium group was higher than that of the other two groups-as aging is associated with a dampening of circadian processes^7,48,59^. Finally, the generalizability of the findings is limited. Because the cohort consisted exclusively of critically ill patients with COVID-19, it is unclear whether similar patterns would be observed in other ICU populations or clinical settings. Further validation in diverse patient cohorts is necessary to assess broader applicability.

Together, these findings demonstrate that wavelet-based analysis offers a powerful and interpretable framework for uncovering dynamic, non-stationary physiological rhythms in routine ICU data and for linking their disruption to adverse outcomes. By providing time-resolved rhythmicity measures that go beyond stationary assumptions, this approach has the potential to enhance continuous monitoring, support earlier detection of deterioration or delirium, and guide rhythm-aware interventions in the ICU. Future work should validate these findings in broader ICU populations and evaluate the integration of wavelet-derived indices into real-time clinical decision-support systems.

## Methods

### Study participants and registration

We included ICU patients treated at Charité – Universitätsmedizin Berlin between January 2020 and February 2022 who were aged ≥ 18. All included patients had an episode of Coronavirus disease 2019 (COVID-19) confirmed by a positive Reverse Transcription Polymerase Chain Reaction (RT-PCR) test for SARS-CoV-2 and required hospitalization. Patients were eligible if they received at least five consecutive days of ICU care. For individuals with multiple hospital admissions, the longest stay was selected for analysis.

The study was approved by the ethics committee of the Charité – Universitätsmedizin Berlin and has been registered in the German Clinical Trials Register (DRKS00036535). The requirement for informed consent was waived due to the retrospective design and use of pseudonymized data. We adhere to the RECORD guidelines for reporting^60^. A CONSORT diagram can be seen in Figure 4.

### Data selection

Data were extracted from Charité’s central research data lake^61^. Access to pseudonymized electronic health records was granted following ethical approval and a data protection impact assessment. Measurement of vitals such as HR and MBP for a continuous five-day period was extracted. The vitals had a measurement point every 30 minutes. The standard data retrieval period was five consecutive days of HR and MBP measurement per patient, selecting days only when delirium or sedation status was simultaneously available. However, to assess the impact of the CoI on signal length, 15 days of vital data were extracted for one subject. Additionally, a key processing rule involved retaining a day with at least five measurements and with intervals between vital measurements not more than four hours. Outlier measurements, defined as those more than three standard deviations from the patient-specific average, were removed. We excluded days that require imputation of over 12 hours or more. Data resampling was performed by segmenting the timeline into 30-minute windows and by taking the median of measurements that fell within each window, leading to a consistent time series. Missing vital measurements were imputed using forward and backward filling methods.

Only patients with five continuous measurement days within the first three weeks post-admission were considered. Since local timestamps in the European/Berlin time zone were retained, the timestamps are prone to daylight saving time changes. Therefore, patients treated during daylight changes might miss or have a few additional timestamps.

### Clinical variables

Daily values of key clinical scores were aggregated to represent the worst clinical status observed on each day. The maximum value was chosen for all the scores except the Glasgow Coma Scale (GCS). For the Richmond Agitation Sedation Scale (RASS), both the minimum and maximum values within each day were retrieved. Following this, an overall score was determined by averaging over five measurement days. Whereas the minimum recorded GCS was selected (indicating greater impairment) on the day of hospital discharge.

The disease/condition definitions were based on the first three characters (category level) of the ICD-10 codes, thereby allowing broad categorization of medical diagnoses (Supplementary Table 3). The generic name of the medications was considered to ensure consistent identification, and the duration of relevant medication was calculated as the drug was administered within the five measurement days, irrespective of its initiation or termination outside this period. A minimum duration of a minute was chosen to represent medication administered at a “point-in-time”. Data for the table were compiled and generated using the tableone Python package (v0.9.5)^62^. Categorical variables are presented as n (%) and continuous variables, except the duration of medications, are shown as median [first quartile (Q1), third quartile (Q3)]. Medication data are reported as mean [minimum, maximum]. Depending upon the variable, the numerical value was rounded to one decimal place or the nearest whole number.

### Outcome definition

Delirium was defined based on the positive Confusion Assessment Method for the ICU (CAM-ICU) or manual review of the physician summary within the patient’s chart showing descriptions of delirium, such as drowsy, confused, disoriented, agitated, and delirious. The aggregation of day-wise delirium status was done by defining a positive status when one positive CAM-ICU entry was recorded for the day. Deep sedation was defined as a day when all documented RASS scores were below -2, indicating moderate or deep sedation (Figure 7A). Based on the delirium or sedation data collected daily across five measurement days, groups were categorized as illustrated in Figure 7B.

**Figure 7.**
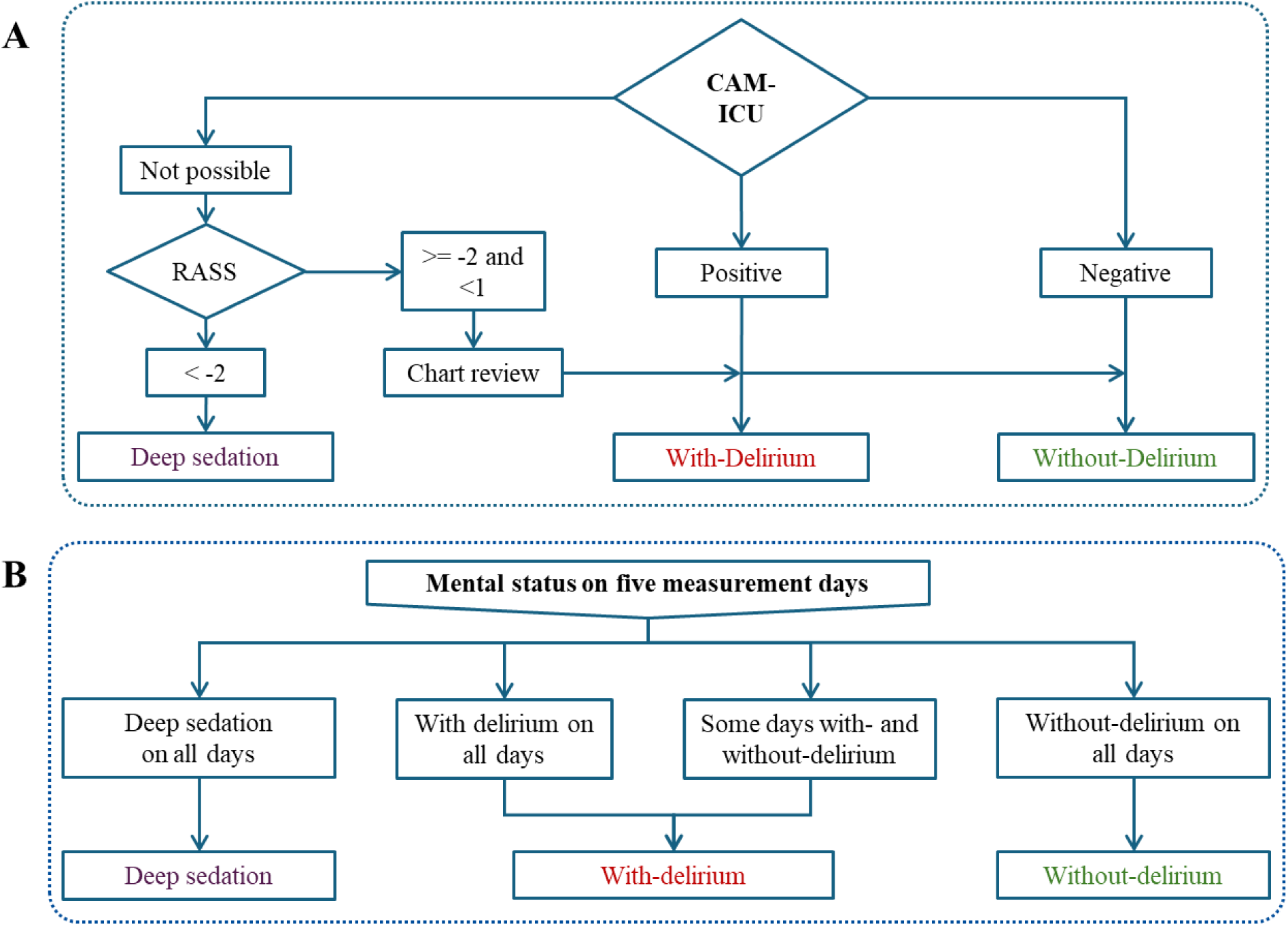
Definition of study groups based on delirium or sedation status. (A) Patients’ delirium or sedation status was determined either by using CAM-ICU or RASS or chart review. (B) Based on delirium or sedation status on five continuous measurement days, patients were categorized into three distinct groups: Deep sedation, With-delirium, Without-delirium. Abbreviations: CAM-ICU, Confusion Assessment Method for the ICU; RASS: Richmond Agitation Sedation Scale

### Wavelet analysis

CWT was implemented with pyBOAT (v0.9.12)^63,64^. To estimate the instantaneous period with CWT, a sequence of 250 evenly spaced values between and including 2 and 48 was generated. The power spectrum of a signal, also called a spectrogram, was generated after applying CWT to the signals. As the CWT yields edge effects due to the finite extent of the wavelets and the convolution operation, the CoI was marked on the spectrogram using white dashed lines.

The amplitude envelope of the Morlet wavelet is given by^65^

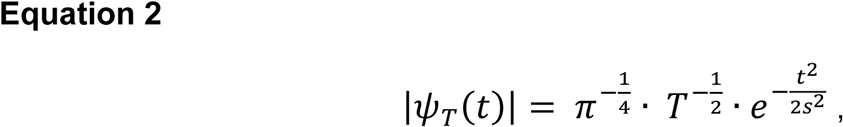

where t represents time and T the period. The CoI is typically delineated by demanding the amplitude envelope decay to 1/𝑒.

### Multi-peak identification, tracking, and segmentation

Bright regions in the CWT power spectrum denote periodic components within the signal and provide direct visualization of their period, magnitude, and temporal localization. To characterize the multiple oscillations, we extended the conventional ridge identification method, which is limited to identifying the maximum power wavelet spectrum, to capture multiple oscillatory components. To this end, we used the find_peaks() function from the scipy package (v1.14.1)^66^ on each frame of the spectrogram. We determine all peaks (dominant and non-dominant oscillations) in the power spectrum by considering a power threshold of 3 to avoid picking up noise in the spectrum.

Our approach for peak tracking uses the Euclidean distance metric to separate individual periodic oscillations. We based this approach on the oscillation period across time. We calculated the Euclidean distance between maxima in consecutive frames (Equation 3), that is, the period of the peaks (T(t), T(t+1)), and recorded which two data points have the minimum distance.

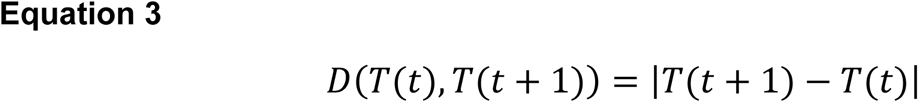

where 𝑇(𝑡) represents the instantaneous period at time 𝑡 and 𝑇(𝑡 + 1) represents the instantaneous period at time 𝑡 + 1.

Periodic components were segmented during iteration based on two criteria: (1) when the Euclidean distance between consecutive data points was greater than or equal to 2, the periodic component either began or ended; and (2) the presence of additional data points during comparison was treated as the origin of a new periodic component. This allowed us to distinguish persistent oscillations from transient oscillations.

### Cosinor Model

All cosinor models have the form

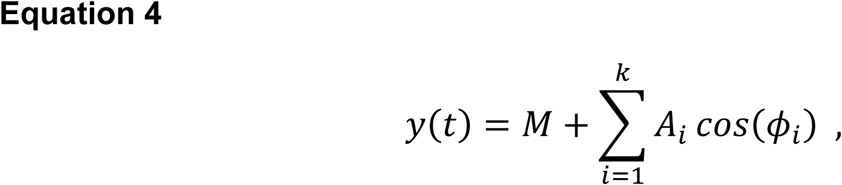

where 𝑦(𝑡) represents HR value at time 𝑡, 𝑀 is the mesor (midline estimating statistic of rhythm) optimized using the least-squares fit, 𝐴_𝑖_ and 𝜙_𝑖_ are the amplitude and phase of the periodic components.

The traditional single- and two-component cosinor models are given for 𝑘 = 1 and 𝑘 = 2, with the instantaneous phase being defined as

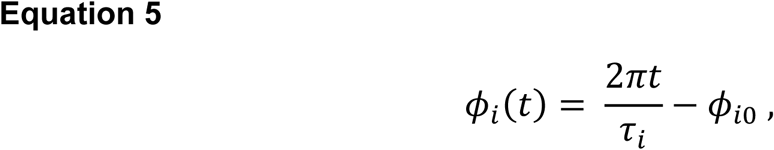

where 𝑡 represents time, 𝜏_𝑖_ the period and 𝜙_𝑖0_ the constant phase offset of the i-th component.

Parameters were optimized using the non-linear least-squares minimization lmfit library (v1.3.4)^67^. To mitigate potential edge effects, the analysis was restricted to periodic oscillations falling outside the CoI. The temporal boundaries were defined by the start and end points of the highest-period oscillation within the region unaffected by the edge effects; as such, the cosinor fit was applied to the data within this window.

For the dynamic modelling approach, we only fitted the mesor (𝑀) and read instantaneous amplitude (𝐴_𝑖_) and phase (𝜙_𝑖_) of the segmented periodic components.

The goodness of fit for the one-component, two-component, and wavelet-derived models was evaluated using the residual sum of squares (R^2^).

### Cohort-specific group heterogeneity

We obtained spectrograms for each patient. Then each spectrogram was first normalized by the sum of all its spectral values. The group spectrum was then calculated as the average of these normalized spectrograms across all patients in the group, to ensure that each individual is weighted equally, independent of absolute power.

Having derived the period, power, and time of the peaks from the spectrum, we obtained a bi-dimensional histogram using the histogram2d function from the numpy library (v1.26.4)^68^. Arbitrary bin widths were defined for each signal component to capture subtle differences between the groups: period (numpy.arange(2, 50, 2)), power (numpy.arange(0, 41, 5)), and time (numpy.arange(0, 120, 4)). We normalized each bin’s raw count by the sum of all bin counts in the histogram.

### Summarization of segmented periodic oscillations

Oscillation characteristics were derived from periodic oscillations situated outside the CoI and were calculated exclusively for periodic components within specific period bands (10-14 and 22-26). Descriptive statistics for continuous variables are reported as median [Q1, Q3], and categorical variables as count (percentage). Differences between groups were assessed using the Kruskal-Wallis test for continuous parameters and the Chi-squared test for categorical parameters. The p-values were adjusted using the Bonferroni correction for each set of comparisons, where the correction factor was determined by the number of tests per set (n=14); an adjusted p-value < 0.05 was considered statistically significant. The tabulation was implemented using the TableOne package (v0.9.5)^62^ in Python.

## Data availability

The data that supports the findings of this study is restricted by ethical limitations and so is not publicly available. Any reasonable request should be made to any of the corresponding authors of this paper.

## Code availability

To enable reproducibility, the complete and annotated code is available online on GitHub: https://github.com/Jayanth-Sreekanth/CWT-HR_MBP-Delir_deepsed.

## Supporting information

RECORD Checklist

## Acknowledgments

This research was initially supported by the Will Foundation (AnäVPostDelir – 137017) and subsequently funded by the Deutsche Forschungsgemeinschaft (DFG, German Research Foundation) – Project-ID 541063275 – TRR 418.

This research has been conducted using data obtained from Charité’s Health Data Platform (HDP)/Medical Data Integration Center (MeDIC). The authors would like to express their sincere gratitude to Denis Beltrami, Maike Lucia Lyall, Annett Roemer, Dr. med. Franziska Lezius for their generous clinical support during data wrangling and curation.

## Author Contributions

S.D.B and J.S. conceptualized the study and developed the methodology. J.S. and A.E. managed data acquisition and curation. J.S. conducted the analysis. S.D.B, J.S., E.S.B., E.G., A.E., and C.S. interpreted the results. E.S.B., A.E. and C.S. provided clinical expertise and medical interpretation of the findings. S.K.P. provided guidance on statistical methods. C.S. and F.B. acquired the funding. J.S. and S.D.B wrote the manuscript, with critical review from all other authors. All authors reviewed and approved the final manuscript.

## Competing interests

J.S., E.S.B. and E.G. report no conflict of interest with respect to the study. A.E. declares that his institution received payments or honoraria for lectures and expert testimony from Gilead Sciences GmbH and Dräger Medical Deutschland GmbH. He holds shares of BioNTech and Novavax (<5000€). A.E. declares to be a member of the German Society of Anaesthesiology and Intensive Care Medicine (DGAI) and participates in the DGAI Round Table on Tele-ICU. Additionally, A.E. collaborates in the section on Metabolism and Nutrition of the German Interdisciplinary Association for Intensive and Emergency Medicine (DIVI). He holds memberships in several other organizations, including the German Sepsis Society (DSG), the European Society of Intensive Care Medicine (ESICM), the European Society of Anaesthesiology and Intensive Care (ESAIC), the German Society for Nutritional Medicine (DGEM), and the European Society for Clinical Nutrition and Metabolism (ESPEN), although he is not actively involved in committee work in these organizations. S.K.P. has no conflicts of interest with respect to the study. Outside the submitted work, she has participated in several Data Safety Monitoring Board meetings that are completely unrelated to this work and the co-authors. C.S. reports grants from the German Research Society (Deutsche Forschungsgemeinschaft), Einstein Foundation Berlin, German Federal Joint Committee (Gemeinsamer Bundesausschuss G-BA), Inner University Grants, Stifterverband (non-profit society promoting science and education), European Society of Anaesthesiology and Intensive Care, German Federal Ministry for Economic Affairs and Energy (BMWE), Georg Thieme Verlag, Dr. F. Köhler Chemie GmbH, Sintetica GmbH, Stifterverband für die Deutsche Wissenschaft e.V. (Medtronic and Teladoc Health), Philips Electronics Nederland BV, German Federal Ministry of Research, Technology and Space (BMFTR), Robert Koch Institute - a Federal Institute within the portfolio of the German Federal Ministry of Health, and the European Commission Horizon Europe during the conduct of the study. In addition, C.S. has different patents and an unpaid leadership or fiduciary role in the German Research Foundation (Deutsche Forschungsgemeinschaft), German National Academy of Sciences–Leopoldina (Deutsche Akademie der Naturforscher Leopoldina e. V.), Berlin Medical Society (Berliner Medizinische Gesellschaft), ESICM (European Society of Intensive Care Medicine), ESAIC (European Society of Anaesthesiology and Intensive Care), German Society of Anaesthesiology and Intensive Care Medicine (DGAI), German Interdisciplinary Association for Intensive Care and Emergency Medicine (DIVI) and German Sepsis Foundation (Deutsche Sepsis-Stiftung). C.S. has participated on the Data Safety Monitoring Board or Advisory Board of Takeda Pharmaceutical Company Limited, Lynx Health Science GmbH and Mundipharma. F.B. reports grants from the German Research Society (Deutsche Forschungsgemeinschaft), Berlin Institute of Health, Berlin University Alliance, Brandenburg Medical School Theodor Fontane, Charité Foundation, Einstein Foundation, European Commission, Federal office for Information Security (BSI), Fresenius Foundation, German Federal Ministry of Education and Research (BMBF), German Federal Ministry of Health (BMG), German Federal Ministry of Research, Technology and Space (BMFTR), German Portal for Medical Research Data (FDPG), Hans Boeckler Foundation, Honda Research Foundation, Joint Federal Committee, Masimo and Philips. Outside the submitted work, F.B. reports royalty payment from Elsevier publishing and has also received financial support from Pfizer for presentations. F.B. reports support for attending meetings from the Association of German Anesthetists, Bavarian Academy of Science and Humanities, German Society of Anesthesiology and Intensive Care Medicine, Masimo and Robert Koch Institute. S.D.B declares that he is a salaried employee at Pfizer Pharma GmbH and his employment has no connection to this work. Outside the submitted work, S.D.B reports financial support from DGPharMed e.V. for a lecture. In addition, he holds shares of Pfizer Pharma GmbH unrelated to this work.

## Supplementary Figures and Tables

By applying CWT to a subject’s HR signal across various durations (3, 5, 7 and 15 days - Supplementary Figure 1A, Supplementary Figure 1B, Supplementary Figure 1C, and Supplementary Figure 1D), we quantified the effect of data length on periodic components. This application shows the impact of edge effects and the minimum number of cycles required to reliably track multiple periodic components (diurnal, semi-diurnal). Based on these results, we recommend using at least five cycles of data to accurately characterize 24-h periodic components spanning multiple days (Supplementary Figure 1B); however, this necessitates excluding the first and last segments of the recording to avert edge effects, with the exclusion duration determined by the CoI for the target period.

**Supplementary Figure 1.**
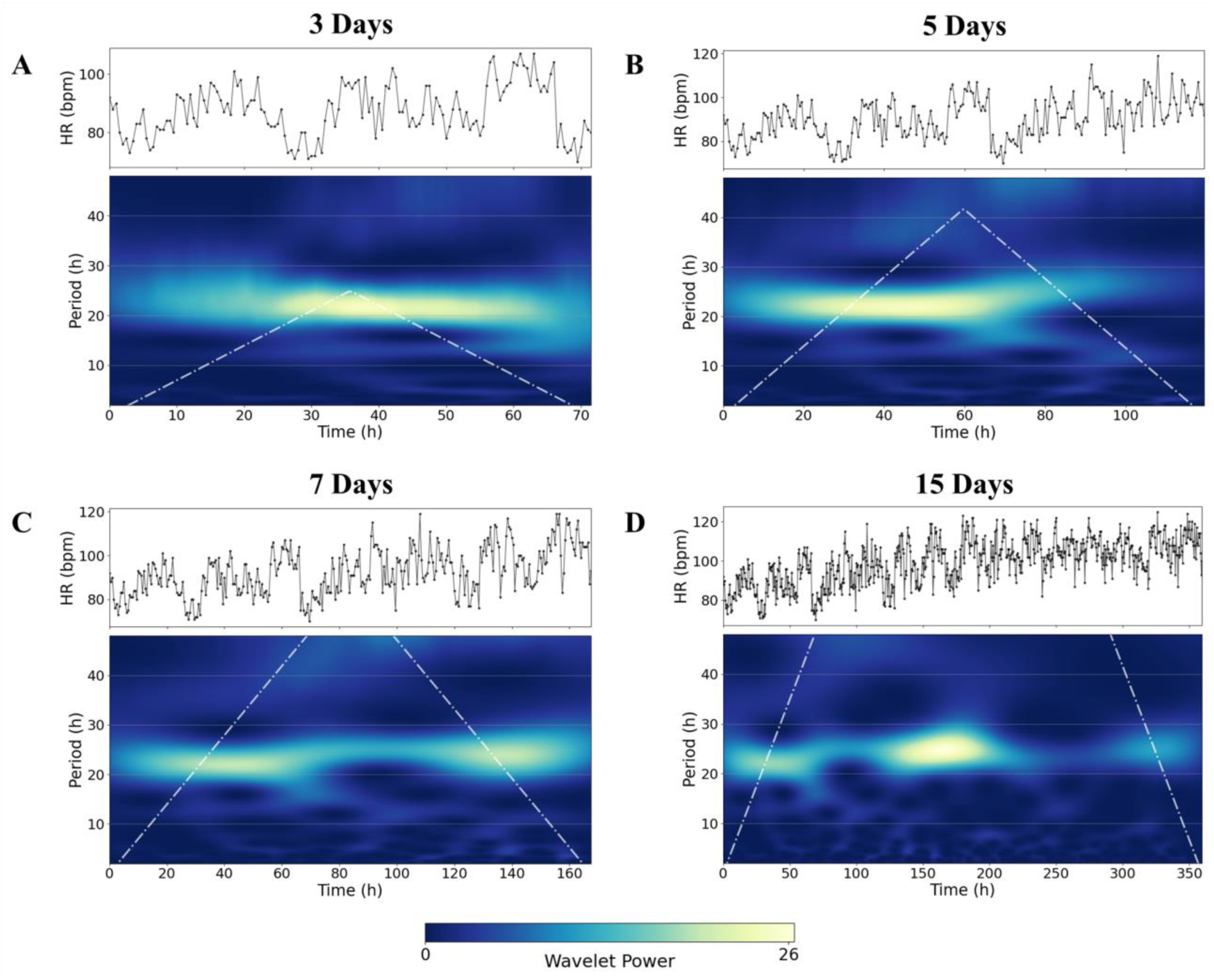
Power spectrum of the CWT for varying signal lengths of the HR signal. The white lines in the spectrograms referring to the CoI can be seen widening as the signal duration increases. A signal with a length of: *(A)* 3-days limits the interpretation of periodic oscillations as the regions with maximum power mostly lie in the area prone to edge effects, *(B)* 5-days shows a good compromise, providing adequate resolution for both diurnal and semi-diurnal oscillations, *(C)* 7-days extends the resolution by enabling the detection of 40-h periodicity, and *(D)* 15-days shows the edge effects captured by CoI have the least impact on the periodic oscillations. Abbreviations: CWT, Continuous Wavelet Transform; HR, Heart Rate; bpm, beats per minute; h, hours; CoI, Cone of Influence

**Supplementary Figure 2.**
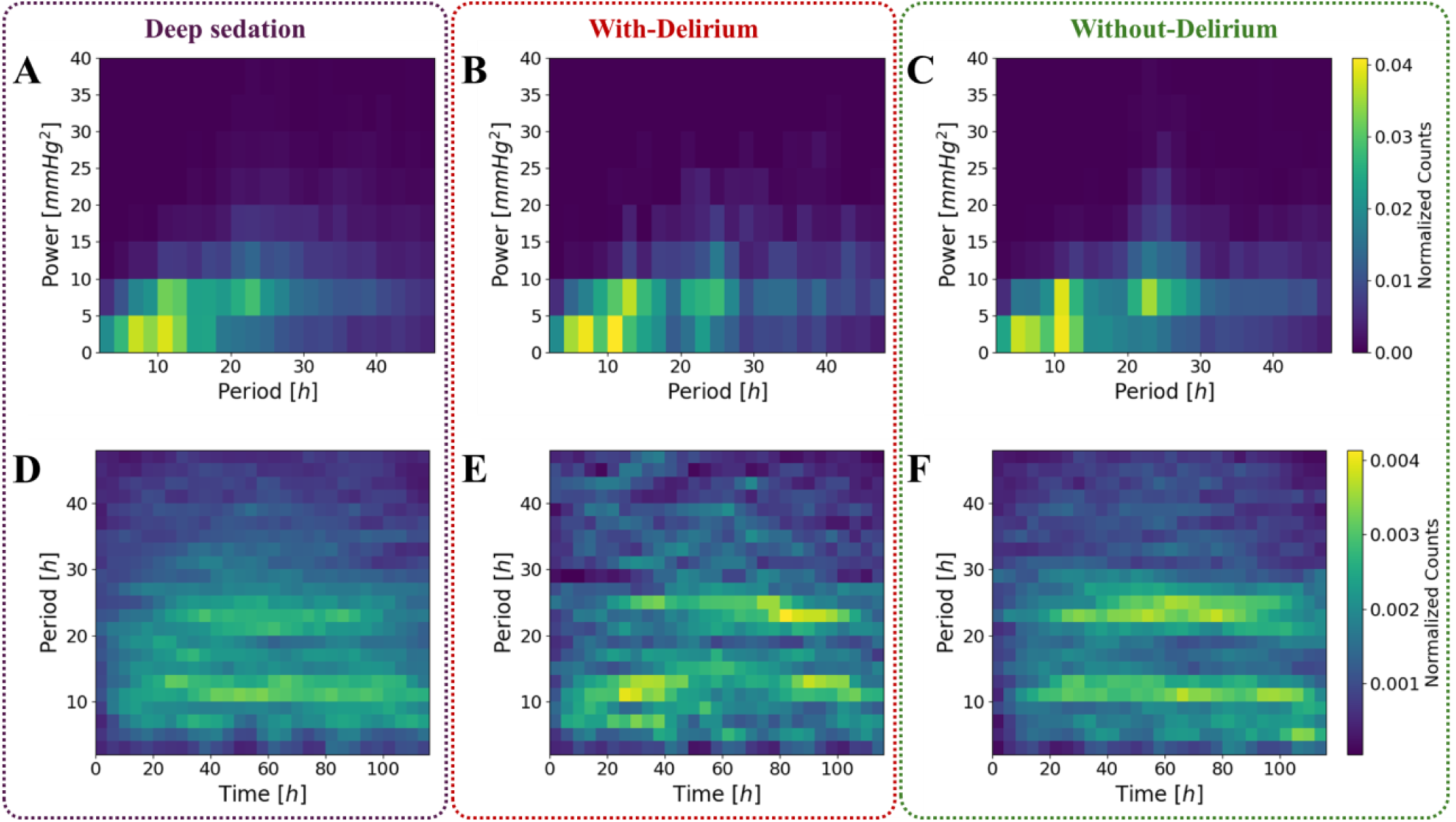
Power spectrum-derived joint distribution of periodic oscillations in MBP measurements. The period in hours (x-axis) and power in mmHg^2^ (y-axis) of oscillations in the *(A)* deep sedation, *(B)* with-delirium, and *(C)* without-delirium group. The trajectory of oscillation period across time in hours in the *(D)* deep sedation, *(E)* with-delirium, and *(F)* without-delirium group are shown. Abbreviations: MBP, Mean Blood Pressure; mmHg, millimeter(s) of mercury; h, hours

**Supplementary Table 1.** An extended summary of the clinical characteristics of the patient cohort. Categorical variables are shown as counts (percentages), while continuous variables are expressed as medians [Q1, Q3].

|  | Missing | Overall | Deep sedation | With-Delirium | Without-Delirium |
| --- | --- | --- | --- | --- | --- |
| n |  | 855 | 439 | 98 | 318 |
| **Respiratory system disorders** |  |  |  |  |  |
| Asthma |  | 50 (5.8) | 29 (6.6) | 2 (2.0) | 19 (6.0) |
| Other chronic obstructive pulmonary disease |  | 65 (7.6) | 29 (6.6) | 9 (9.2) | 27 (8.5) |
| **Metabolic disorders** |  |  |  |  |  |
| Type 1 diabetes mellitus |  | 8 (0.9) | 3 (0.7) | 0 (0.0) | 5 (1.6) |
| Type 2 diabetes mellitus |  | 241 (28.2) | 118 (26.9) | 35 (35.7) | 88 (27.7) |
| Obesity |  | 160 (18.7) | 114 (26.0) | 4 (4.1) | 42 (13.2) |
| Disorders of lipoprotein metabolism and other lipidaemias |  | 137 (16.0) | 34 (7.7) | 33 (33.7) | 70 (22.0) |
| **Musculoskeletal disorders** |  |  |  |  |  |
| Osteoporosis without pathological fracture |  | 2 (0.2) | 0 (0.0) | 0 (0.0) | 2 (0.6) |
| Seropositive rheumatoid arthritis |  | 6 (0.7) | 3 (0.7) | 1 (1.0) | 2 (0.6) |
| Other rheumatoid arthritis |  | 14 (1.6) | 2 (0.5) | 3 (3.1) | 9 (2.8) |
| Gonarthrosis [arthrosis of knee] |  | 1 (0.1) | 0 (0.0) | 0 (0.0) | 1 (0.3) |
| Other arthrosis |  | 2 (0.2) | 1 (0.2) | 0 (0.0) | 1 (0.3) |
| **Digestive system disorders** |  |  |  |  |  |
| Crohn disease [regional enteritis] |  | 3 (0.4) | 1 (0.2) | 0 (0.0) | 2 (0.6) |
| Ulcerative colitis |  | 3 (0.4) | 0 (0.0) | 0 (0.0) | 3 (0.9) |
| **Urogenital disorders** |  |  |  |  |  |
| Chronic kidney disease |  | 111 (13.0) | 38 (8.7) | 31 (31.6) | 42 (13.2) |

**Supplementary Table 2.** Clinical characteristics of the group across five measurement days, including clinical scores and medications stratified according to their type. The variables except medications are reported as median [Q1, Q3] and the duration of medications in minutes is shown as mean [minimum, maximum]. Abbreviations: MBP, Mean Blood Pressure; HR, Heart Rate; RASS, Richmond Agitation-Sedation Scale; CAM-ICU, Confusion Assessment Method for the Intensive Care Unit; Hydromorphone-HC, Hydromorphone Hydrochloride; APACHE II, Acute Physiology and Chronic Health Evaluation II; SOFA, Sequential Organ Failure Assessment; TISS-10, Therapeutic Intervention Scoring System-10; Q1, first quartile; Q3, third quartile

|  | Missing | Overall | Deep sedation | With-Delirium | Without-Delirium |
| --- | --- | --- | --- | --- | --- |
| **n** |  | 855 | 439 | 98 | 318 |
| Number of patients required imputation for MBP and HR |  | 218 (25.5) | 79 (18.0) | 27 (27.6) | 112 (35.2) |
| Number of imputed timestamps for MBP and HR | 637 | 10 [4,20] | 8 [4,19] | 5 [2,13] | 11 [6,21] |
| minimum RASS | 0 | -3 [-5,0] | -5 [-5,-4] | -1 [-5,0] | 0 [-2,0] |
| maximum RASS | 0 | -2 [-5,3] | -4 [-5,-3] | 1 [-1,3] | 0 [-1,1] |
| Days with "positive" CAM-ICU | 0 | 0 [0,0] | 0 [0,0] | 1 [1,2] | 0 [0,0] |
| Days with "unknown" CAM-ICU | 0 | 4 [0,5] | 5 [5,5] | 0 [0,1] | 0 [0,0] |
| Days with "negative" CAM-ICU | 0 | 0 [0,5] | 0 [0,0] | 2 [1,4] | 5 [5,5] |
| **Analgesic Medications (duration in minutes)** |  |  |  |  |  |
| Buprenorphine | 0 | 0 [0,7] | 0 [0,0] | 0 [0,7] | 0 [0,0] |
| Fentanyl | 0 | 0 [0,8] | 0 [0,3] | 0 [0,8] | 0 [0,8] |
| Hydromorphone-HC | 0 | 0 [0,8] | 0 [0,0] | 0 [0,6] | 0 [0,8] |
| Morphine | 0 | 7 [0,5761] | 13 [0,5761] | 0 [0,2] | 0 [0,6] |
| Oxycodone+Naloxone | 0 | 0 [0,8] | 0 [0,0] | 0 [0,6] | 0 [0,8] |
| Pethidine | 0 | 0 [0,60] | 0 [0,1] | 0 [0,0] | 0 [0,60] |
| Piritramide | 0 | 0 [0,13] | 0 [0,0] | 0 [0,7] | 0 [0,13] |
| Ropivacaine + Sufentanil | 0 | 1 [0,1047] | 0 [0,0] | 11 [0,1047] | 0 [0,0] |
| Sufentanil | 0 | 125 [0,5867] | 231 [0,5867] | 55 [0,5161] | 2 [0,615] |
| Tramadol | 0 | 0 [0,8] | 0 [0,0] | 0 [0,7] | 0 [0,8] |
| **Sedatives (duration in minutes)** |  |  |  |  |  |
| Clonidine/<br>Dexmedetomidine | 0 | 0 [0,3] | 0 [0,0] | 0 [0,3] | 0 [0,1] |
| Esketamine | 0 | 65 [0,6289] | 126<br>[0,6289] | 0 [0,1] | 0 [0,0] |
| Ketamine | 0 | 129<br>[0,5764] | 251<br>[0,5764] | 0 [0,0] | 0 [0,1] |
| Lorazepam | 0 | 0 [0,3] | 0 [0,0] | 0 [0,2] | 0 [0,3] |
| Midazolam | 0 | 30 [0,5763] | 58 [0,5763] | 0 [0,2] | 0 [0,0] |
| Propofol | 0 | 163<br>[0,5952] | 283<br>[0,5952] | 78 [0,5358] | 24<br>[0,3574] |
| **Vasopressors (duration in minutes)** |  |  |  |  |  |
| Epinephrine | 0 | 21 [0,5758] | 31 [0,5758] | 2 [0,108] | 12<br>[0,3783] |
| Norepinephrine | 0 | 1014<br>[0,8041] | 1772<br>[0,8041] | 395<br>[0,4942] | 159<br>[0,5123] |
| **Scores** |  |  |  |  |  |
| APACHE II | 271 | 26 [16,32] | 31 [27,36] | 20 [17,24] | 13<br>[10,17] |
| SOFA | 1 | 8 [3,11] | 11 [10,12] | 4 [3,6] | 2 [1,3] |
| TISS-10 | 0 | 14 [10,17] | 17 [15,20] | 10 [8,13] | 10 [8,12] |

**Supplementary Table 3.** Categorization of disorders and diseases according to ICD-10-GM (International Statistical Classification of Diseases and Related Health Problems version 10 with German Modification) standards. * represents that all succeeding codes are classified under the listed category.

| **Disorder and Disease** | **Category** | **Code** |
| --- | --- | --- |
| *Circulation system disorders* | Atherosclerosis | I70.* |
|  | Chronic ischaemic heart disease | I25.* |
|  | Heart failure | I50.* |
|  | Essential (primary) hypertension | I10.* |
|  | Hypertensive heart disease | I11.* |
|  | Hypertensive renal disease | I12.* |
|  | Hypertensive heart and renal disease | I13.* |
| *Respiratory system disorders* | Asthma | J45.* |
|  | Other chronic obstructive pulmonary disease | J44.* |
| *Metabolic disorders* | Type 1 diabetes mellitus | E10.* |
|  | Type 2 diabetes mellitus | E11.* |
|  | Obesity | E66.* |
|  | Disorders of lipoprotein metabolism and other lipidaemias | E78.* |
| *Mental and behavioral disorders* | Depressive episode | F32.* |
|  | Recurrent depressive disorder | F33.* |
|  | Bipolar affective disorder | F31.* |
|  | Schizophrenia | F20.* |
|  | Other anxiety disorders | F41.* |
| *Neurological disorders* | Alzheimer disease | G30.* |
|  | Parkinson disease | G20.* |
|  | Multiple sclerosis | G35.* |
| *Musculoskeletal disorders* | Osteoporosis without pathological fracture | M81.* |
|  | Seropositive rheumatoid arthritis | M05.* |
|  | Other rheumatoid arthritis | M06.* |
|  | Gonarthrosis [arthrosis of knee] | M17.* |
|  | Other arthrosis | M19.* |
| *Digestive system disorder* | Crohn disease [regional enteritis] | K50.* |
|  | Ulcerative colitis | K51.* |
| *Urogenital disorder* | Chronic kidney disease | N18.* |

**Supplementary Table 4.** Inter-group differences of the group summarized periodic components in MBP oscillations. All continuous variables are presented as median [Q1, Q3] and categorical variables as count (percentage). Statistical significance was assessed using the Kruskal-Wallis test for continuous variables and the Chi-squared test for categorical variables. P-values were adjusted using the Bonferroni method applied independently to each set of comparisons with a correction factor of n=14. The patients excluded are the ones who didn’t have oscillations within the specific period or had specific periodic oscillations inside the CoI. Abbreviations: MBP, Mean Blood Pressure; h, hours; mmHg, millimeter(s) of mercury; N.S., Not Significant; Q1, first quartile; Q3, third quartile; CoI, Cone of Influence

| Oscillation Characteristics | Oscillation Period (h) | Overall | Deep sedation | With-Delirium | Without-Delirium | P-Value (adjusted) [Deep sedation vs With-Delirium] | P-Value (adjusted) [Deep sedation vs Without-Delirium] | P-Value (adjusted) [With-Delirium vs Without-Delirium] | P-Value (adjusted) [Overall] |
| --- | --- | --- | --- | --- | --- | --- | --- | --- | --- |
| n |  | 855 | 439 | 98 | 318 |  |  |  |  |
| Average duration of oscillations (h) | 10 - 14 | 19 [10,30] | 19 [10,30] | 20 [11,30] | 20 [11,32] | N.S. | N.S. | N.S. | N.S. |
| Minimum oscillation duration (h) |  | 12 [4,27] | 10 [3,26] | 10 [2,26] | 12 [4,30] | N.S. | N.S. | N.S. | N.S. |
| Maximum oscillation duration (h) |  | 25 [14,40] | 24 [14,39] | 26 [14,40] | 24 [14,41] | N.S. | N.S. | N.S. | N.S. |
| Oscillation start hour |  | 27.0 [18.5,51.5] | 29.0 [18.5,52.0] | 23.5 [17.5,58.5] | 27.0 [18.2,51.5] | N.S. | N.S. | N.S. | N.S. |
| Oscillation end hour |  | 93.0 [67.0,101.5] | 93.5 [69.5,101.5] | 99.5 [61.5,102.0] | 90.5 [62.0,102.0] | N.S. | N.S. | N.S. | N.S. |
| Average power (mmHg<sup>2</sup>) |  | 5 [4,7] | 5 [4,6] | 5 [4,7] | 5 [4,6] | N.S. | N.S. | N.S. | N.S. |
| Count of patients excluded (no peaks or all peaks inside Col) |  | 162 (18.9) | 70 (15.9) | 21 (21.4) | 71 (22.3) | N.S. | 0.463 | N.S. | 0.972 |
| Average duration of oscillations (h) | 22 - 26 | 21 [10,38] | 19 [8,36] | 22 [8,36] | 26 [12,44] | N.S. | 0.127 | N.S. | 0.403 |
| Minimum oscillation duration (h) |  | 21 [8,39] | 18 [7,36] | 22 [7,36] | 26 [12,44] | N.S. | 0.108 | N.S. | 0.361 |
| Maximum oscillation duration (h) |  | 22 [12,39] | 20 [10,36] | 24 [8,36] | 26 [12,44] | N.S. | 0.242 | N.S. | 0.694 |
| Oscillation start hour |  | 38.0 [34.5,57.5] | 37.0 [34.5,57.5] | 45.0 [36.0,62.5] | 37.5 [34.5,53.8] | 0.420 | N.S. | 0.238 | 0.749 |
| Oscillation end hour |  | 82.5 [62.8,85.0] | 81.5 [60.0,85.0] | 84.0 [70.8,85.8] | 83.0 [65.5,85.0] | 0.851 | N.S. | N.S. | N.S. |
| Average power (mmHg<sup>2</sup>) |  | 7 [5,12] | 7 [5,12] | 7 [4,10] | 8 [5,12] | 0.819 | N.S. | 0.435 | N.S. |
| Count of patients excluded (no peaks or all peaks inside Col) |  | 336 (39.3) | 178 (40.5) | 39 (39.8) | 119 (37.4) | N.S. | 0.789 | N.S. | N.S. |

## Notes

### Funding Statement

This research was initially supported by the Will Foundation (AnaeVPostDelir: 137017) and subsequently funded by the Deutsche Forschungsgemeinschaft (DFG, German Research Foundation), Project ID: 541063275, TRR 418.

### Author Declarations

The study was approved by the ethics committee of the Charite Universitaetsmedizin Berlin and has been registered in the German Clinical Trials Register (DRKS00036535). The requirement for informed consent was waived due to the retrospective design and use of pseudonymized data.

